# Meta-analysis of the gut microbiome of Parkinson’s disease patients suggests alterations linked to intestinal inflammation

**DOI:** 10.1101/2020.08.10.20171397

**Authors:** Stefano Romano, George M. Savva, Janis R. Bedarf, Ian G. Charles, Falk Hildebrand, Arjan Narbad

## Abstract

The gut microbiota is emerging as an important modulator of neurodegenerative diseases, and accumulating evidence has linked gut microbes to Parkinson’s disease (PD) symptomatology and pathophysiology. PD is often preceded by gastrointestinal symptoms and alterations of the enteric nervous system accompany the disease. Several studies have analyzed the gut microbiome in PD, but a consensus on the features of the PD-specific microbiota is missing. Here, we conduct a meta-analysis re-analyzing 10 currently available 16S microbiome datasets to investigate whether underlying alterations in the gut microbiota of PD patients exist. We found consistent alterations in PD-associated microbiome, which are significant and robust to confounders across studies, although differences in microbiome structure between PD and controls are limited. Enrichment of the genera *Lactobacillus, Akkermansia*, and *Bifidobacterium* and depletion of bacteria belonging to the families Lachnospiraceae and Ruminococcaceae, which are important short-chain fatty acids producers, emerged as the most consistent PD gut microbiome alterations. This dysbiosis might result in a pro-inflammatory status which could explain the recurrent gastrointestinal symptoms affecting PD patients.

## Introduction

Parkinson’s disease (PD) is the second most common neurodegenerative disorder after Alzheimer’s disease^1^. Globally, it has an incidence of 10-50/100,000 person/year and a prevalence of 100-300/100,000 people, and due to the increase in aging population the number of PD patients is expected to double by 2030^1^. PD affects predominantly dopaminergic neurons in the brain, leading to decreased dopamine levels and motor impairments^2^. Its pathological hallmark has long been considered to be the intracellular deposition of aggregated α-synuclein, leading to neuronal cell death and neuro-inflammation^3^. PD is now considered a multi-systemic disease, affecting the central as well as the enteric nervous system (CNS, ENS), resulting in several non-motor symptoms, often including gastroparesis or constipation. Due to the early involvement of the gastrointestinal tract, often preceding motor symptoms for years^4^, changes in gut microbiota composition have been studied in relation to the pathophysiology of PD. The potential role of gut microbiota in PD^3^ and other neurodegenerative diseases^5^ is supported by animal studies^6^, showing that the microbiota can affect α-synucleinopathy as well as neuro-inflammation. Thus, the microbiota is a putative therapeutic target and has the potential for developing diagnostic biomarkers.

PD patients can have increased gut permeability and inflammation^7,8^, and these have been hypothesized to be linked to low gastrointestinal short-chain fatty acids (SCFA) concentrations^9^. SCFA are the end products of bacterial fermentation of dietary components and play a pivotal role in fueling and maintaining the integrity of the colonic epithelium. Low levels of SCFA have been considered to be a consequence of a decreased abundance of SCFA-producing taxa in PD patients^10,11^. To date, more than 20 cohort-studies have investigated the composition of the PD gut microbiota. Over 100 differently abundant taxa between PD patients and controls have been reported^10–19^, and some studies detected association between taxa abundances and disease severity^11,12,16^. Several studies suggested that PD patients have an altered gut microbiota compared to controls, even though findings are often inconsistent and a consensus on the taxa associated with the disease is still lacking. Across most studies, the genus *Akkermansia* and the Verrucomicrobiaceae family have been found to be enriched in PD patients, while bacteria belonging to the Lachnospiraceae family are depleted. On the other hand, various inconsistencies have been found among the different sampling cohorts. For example, the Lactobacillaceae family has been generally detected to be enriched in PD in the Western cohorts but never in Chinese studies^18,20,21^. Similarly, conflicting results have been obtained for bacteria within the Prevotellaceae family. Several studies detected these taxa to be highly depleted in PD patients^16,17,22,23^, compared to controls, whereas others found no differences in abundances^11^ or found these taxa enriched in PD patients^13,20^.

Inconsistencies amongst studies might arise from variations in study designs and methods used for producing and analyzing 16S rRNA gene amplicon data, as well as from the natural variability of the gut microbiota across populations, lifestyles, and diets. To further elucidate the significance of changes in the intestinal microbiota composition in PD and to evaluate its potential as a biomarker for PD risk, diagnosis, and prognosis it is important to perform cross-study comparisons and identify disease-specific alterations. Here, we provide a thorough meta-analysis (pooled re-analysis) of all ten available studies that described the gut microbiome in PD through 16S rRNA amplicon sequencing. We apply a standardized workflow to analyze each study individually and combined different statistical approaches to identify the major changes affecting the gut microbiome of PD patients across sampling cohorts.

## Results

### Study selection

We identified a total of 22 studies that investigated the PD-associated gut microbiome using 16S rRNA gene amplicon sequencing (Supplementary Table 1). Of these, ten made raw sequencing data available and were re-analyzed in our study. These ten studies covered nine different cohorts (one was reported at baseline then at follow-up two years later), across six countries (Table 1). Overall, this resulted in 1,211 samples (Fig 1A) obtained from all case-control studies. Cases were usually selected from clinics local to the investigating teams, were at different stages of the disease, and almost all were using some form of PD medication (Table 1). Controls were typically sampled by convenience from the local population or from families of the PD patients. All studies except one^16^ applied the UK brain bank criteria to define PD. Cases had an average age of between 60 and 70 years in all studies, with controls typically well matched in age. Some studies matched on sex, while for others there were significantly more males in case compared to control groups^12,13^.

**Table 1.** Technical details of the studies included in the meta-analysis. The table reports the following information: study design (a: disease duration at sampling reported as mean and respective standard deviation, and proportion of medicated patients), sample collection, DNA extraction, and sequencing techniques (b: sequencing platform; c: paired-ends vs single-ends). NA indicates that the information was not reported in the original article. RT indicate room temperature. In many studies the proportion of medicated patients was calculated for individual type of drugs. Hence, we could not estimate the total portion of patients undergoing pharmaceutical treatments, and we report here only the type of drugs with the highest proportions of medicated patients.

| Study | Design | Disease duration & medications<sup>a</sup> | Country | Sampling | DNA extr. | 16S Region | Seq. Tech.<sup>b</sup> | PE vs SE<sup>c</sup> |
| --- | --- | --- | --- | --- | --- | --- | --- | --- |
| Hill-Burns et al.<sup>12</sup> | Case-control | 13.7 ± 6.5 years, 93.9% medicated | USA | Swabs, delivered at RT | MO BIO's PowerMag Soil kit | V4 | MiSeq | SE |
| Hopfner et al.<sup>14</sup> | Case-control | 11.2 ± 4.8 years, all medicated | Germany | Home or Hospital collection, delivered at RT | Power Soil Kit | V1-V2 | MiSeq | PE |
| Keshavarzian et al.<sup>11</sup> | Case-control | 29.7 ± 11 years, 66.4% medicated | USA | Home collection, delivered in GasPak | FastDNA Soil Kit | V4 | MiSeq | SE |
| Petrov et al.<sup>17</sup> | Case-control | NA | Russia | NA | Mechanical disruption | V3-V4 | MiSeq | SE |
| Pietrucci et al.<sup>19</sup> | Case-control | 8.1 ± 4.5 years 82.5% on l-dopa | Italy | Home collection, DNA stabilizer | PSP-Spin Stool Kit | V3-V4 | MiSeq | PE |
| Qian et al.<sup>18</sup> | Matched case-control | 5.7 ± 4.1 years, all medicated | China | Home collection, transferred in Ice | QIAm DNA stool Mini Kit | V3-V4 | MiSeq | PE |
| Aho et al.<sup>16</sup> | Matched case-control | Median 8.5 years, 73% on l-dopa | Finland | Home collection, DNA stabilizer, stored in fridge | PSP-Spin Stool Kit | V3-V4 | MiSeq | PE |
| Scheperjans et al.<sup>15</sup> | Matched case-control | Median 6.5 years, all but 2 using medications (77.8% on dopamine agonist) | Finland | Home collection, DNA stabilizer, stored in fridge | PSP-Spin Stool Kit | V3-V4 | MiSeq | PE |
| Heintz-Buschart et al.<sup>13</sup> | Case-control | 72 ± 31 months, 85% on l-dopa agonist | Germany | Home collection, flash-frozen | Modified Qiagen Allprep | V4 | MiSeq | PE |
| Weis et al.<sup>24</sup> | Matched case-control | 82 ± 56 months, 85.3% medicated | Germany | MED AUXIL fecal collector set | FastDNA Spin Kit | V4-V5 | IonTorrent | PE |

**Figure 1.**
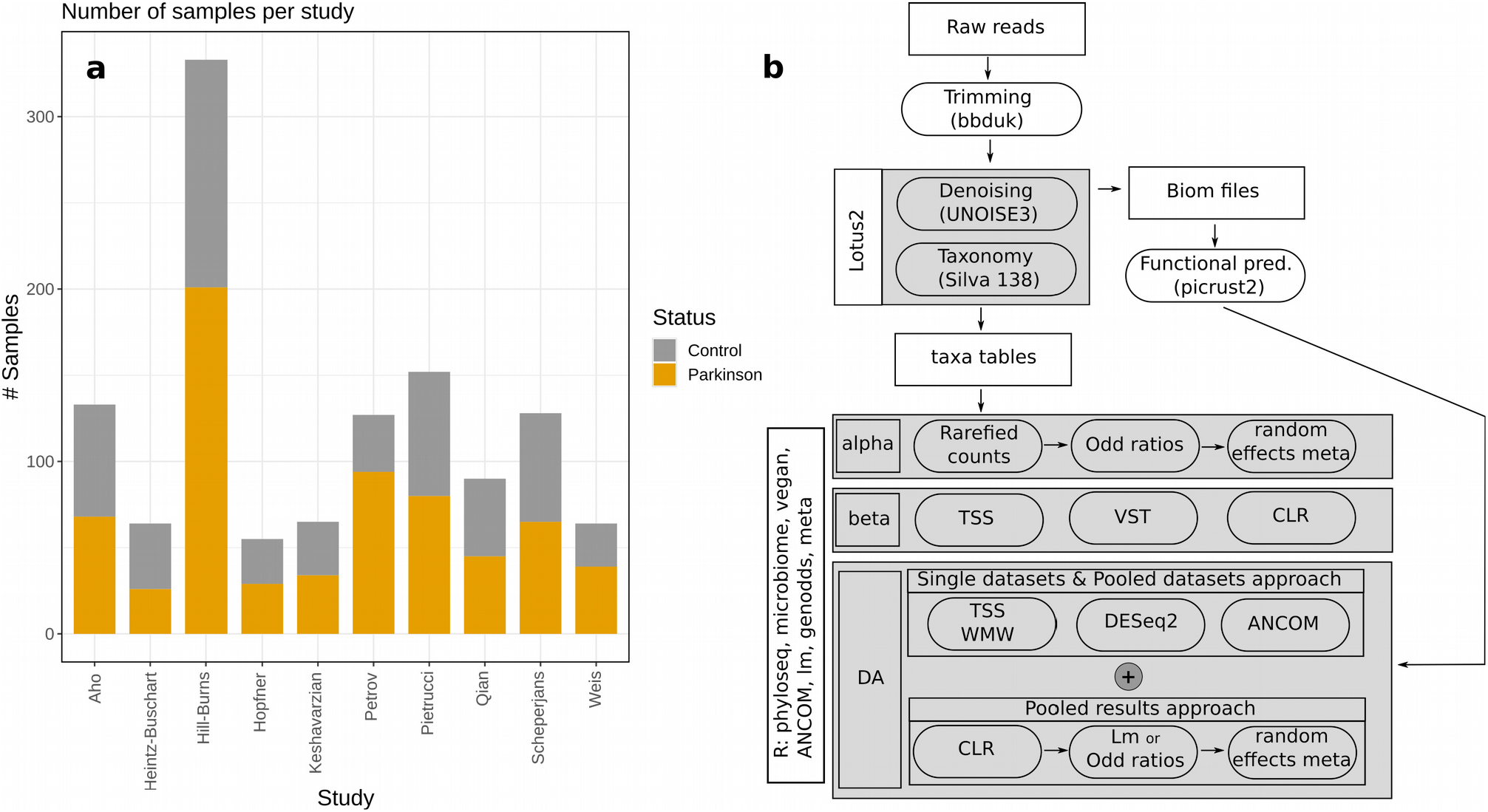
Sample distribution across studies (a) and bioinformatic work-flow adopted in our study (b). The number of control and PD samples is reported and refers to the data that could be recovered from the Sequence Read Archive (SRA) or the European Nucleotide Archive (ENA). TSS = Total Sum Scaling; VST = Variance Stabilizing Transformation; CLR = Centered Log-Ratios; WMW = Wilcoxon-Mann-Whitney test; ANCOM = ANalysis of COmposition of Microbiomes; Lm = linear models.

Various sampling protocols were used across studies, with considerable variation in the method used to preserve the samples before processing (Table 1). In some cases, samples were kept at room temperature for up to 48 hours before analysis^14^, in others, samples were stored either in DNA preservative^16,19^ or on ice^18^. DNA extractions and sequencing strategies also varied across studies (Table 1). The Illumina MiSeq platform was the most used sequencing technology, but the 16S variable region and sequencing strategy (paired-ends vs single-ends) varied considerably (Table 1). Considering the heterogeneity across studies, we first re-analyzed each single dataset individually, then we used a combination of statistical approaches to obtain a consensus overview of gut microbiome structure in PD accounting for the heterogeneity between studies. Two studies were based on the same cohort measured at different time points, hence, we performed a sensitivity analysis by comparing the results obtained considering both datasets with those obtained after omitting the baseline samples^15^.

### The gut microbiome differs significantly between PD patients and controls

Measures of microbial alpha-diversity, based on species profiles, were higher in PD samples compared to controls in three out of ten studies (Supplementary Figure 1). Interestingly, these three datasets were the only ones using single-end sequencing approaches, suggesting that this might influence the estimation of bacterial diversity. These differences were still overall significant when we pooled estimates across studies using random-effects meta-analysis (Fig 2, Supplementary Figure 1). Specifically, PD samples had a higher overall richness as indicated by a significantly higher number of observed species and higher Fisher’s alpha, ACE, and Chao1 indices (Fig 2; Supplementary Figure 1). Our analyses suggest that this higher diversity might derive from a decrease in the abundance of dominant species and an increase in rare/low abundant ones, as dominance indices were lower and rarity indices were significantly higher in PD patients (Fig 2; Supplementary Figure 1). Previous studies reported a higher abundance of Firmicutes in control samples compared to PD^11^, and the Firmicutes to Bacteroidetes ratio (F/B ratio) has been frequently used to assess gut-health. Therefore, we calculated F/B ratios across all studies. Only in the study of Keshavarzian et al^11^ did we observe a significant difference in the F/B ratio between PD and control, but this difference was not overall significant (Supplementary Figure 2). Similarly, Aho et al^16^ reported that controls had an increased *Prevotella* to *Bacteroides* ratio (P/B ratio) in the baseline samples^15^ of their longitudinal study. We confirmed this result and detected an increased P/B ratio in two other studies, but did not detect the same association in the rest of the datasets, and there was only weak evidence for a higher P/B ratio in controls when results were pooled (Supplementary Figure 2). Omitting the baseline samples from the longitudinal Finnish cohort did not alter the conclusions of the alpha-diversity analyses, and led to a decrease in the P/B ratio difference between PD and controls (data not shown).

**Figure 2.**
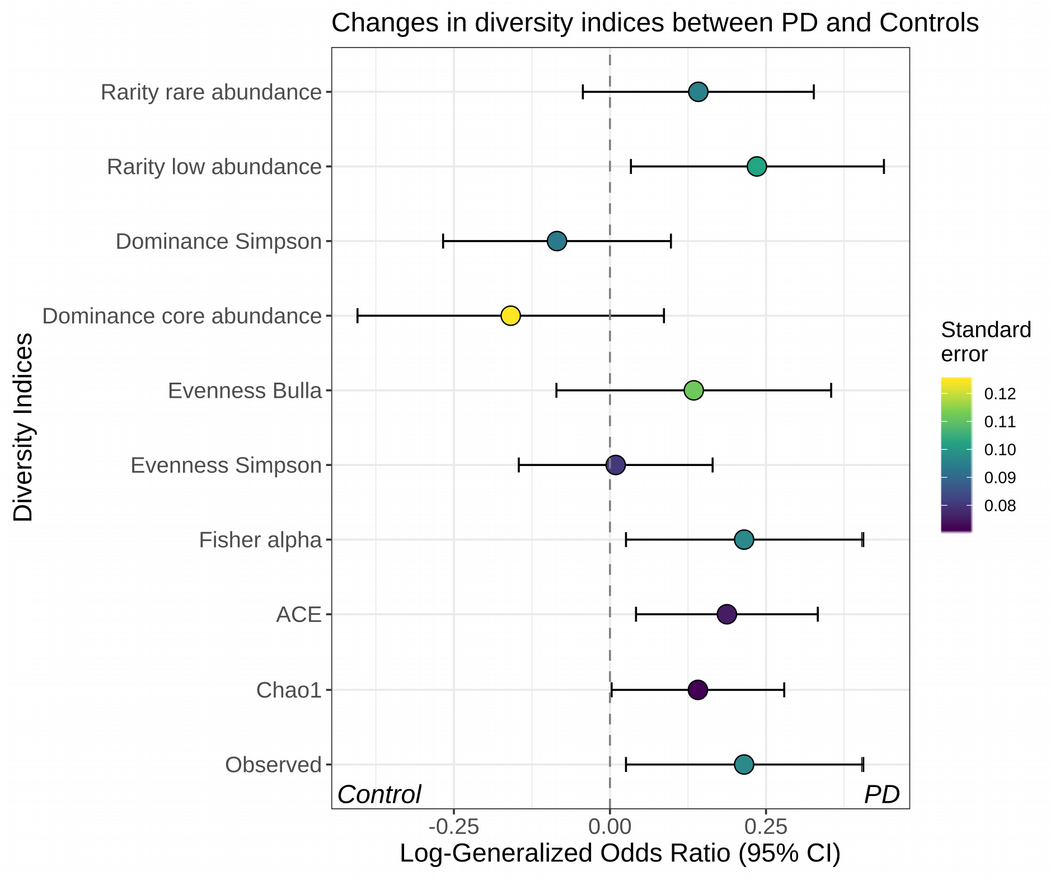
Alpha diversity indices are significantly different between PD patients and controls. Indices were calculated at the species level for each dataset. Results were then combined using a random-effects meta-analysis approach. The log-generalised Odds Ratios indicate the degree of variation of each index between controls and PD. The richness of the samples was estimated using the observed number of species and the Chao1, ACE, and Fisher’s alpha indices. To estimate eveness, which indicates how different the abundances of the species in a community are from each other, we used the Bulla and Simpson indices. Finally, we estimated dominance, which describes how much one or few species dominate the community, and rarity, which assesses the number of species with low abundance in the samples. The data suggest that the gut-microbiota of PD patients is more diverse (higher richness) than controls and this is likely a consequence of an increase in rare taxa (rarity).

The genera *Bacteroides* and *Prevotella* and the Firmicutes phylum are key gut microbiota taxa that have different abundances in the three proposed enterotypes (ET_B, ET_P, and ET_F, representing the *Bacteroides, Prevotella* and Firmicutes enterotypes, respectively)^25^. To verify the prevalence of these gut microbiome types amongst PD patients, we assigned each microbiome to one of the three known enterotypes, and managed to classify 589 samples. The distribution of assigned enterotypes varied enormously across studies, but there was no significant difference between PD and control samples in any individual study and no trend toward specific enterotypes when studies were considered together (Supplementary Figure 3). For example, the majority of PD samples from the Finnish cohort^15,16^ were assigned to the ET_F, whereas in the study of Hill-Burns et al.^12^ and Pietrucci et al.^19^ most of the PD samples were classified as ET_B. Interestingly, only in the Finnish cohorts was the ET_P more common among control samples, in agreement with the authors’ finding of *Prevotella* being enriched in the control groups.

Considering the variability among studies and the potential data-dependent effect of different microbiome analysis workflows^26^, we used a thorough and comprehensive approach to investigate the structure of the bacterial communities associated with PD (Fig 1B). We used three independent normalization strategies (Variance Stabilizing Transformation, VST; Total Sum Scaling, TSS, Centered Log-Ratio, CLR) combined as appropriate with three beta-diversity distances (Bray-Curtis, BC; Jensen-Shannon divergence, JSD; Euclidean) and statistical testing via permutational multivariate analysis of variance using distance matrices (PERMANOVA). We applied these strategies to all three taxonomy ranks we considered (species, genus, and family). In most studies, irrespective of the normalization-distance combinations, disease status significantly explained the differences within the data (p < 0.05), even though it accounted only for a limited portion of data variability (from < 1% to <13%; R^2^ expressed in percentage; Fig 3). When the datasets were pooled, both study and disease status significantly explained the separation of the samples, but the proportion of variance explained by the disease status was in all cases < 1% (Fig 3), whereas the study explained between 28 and 54% of the variance.

**Figure 3.**
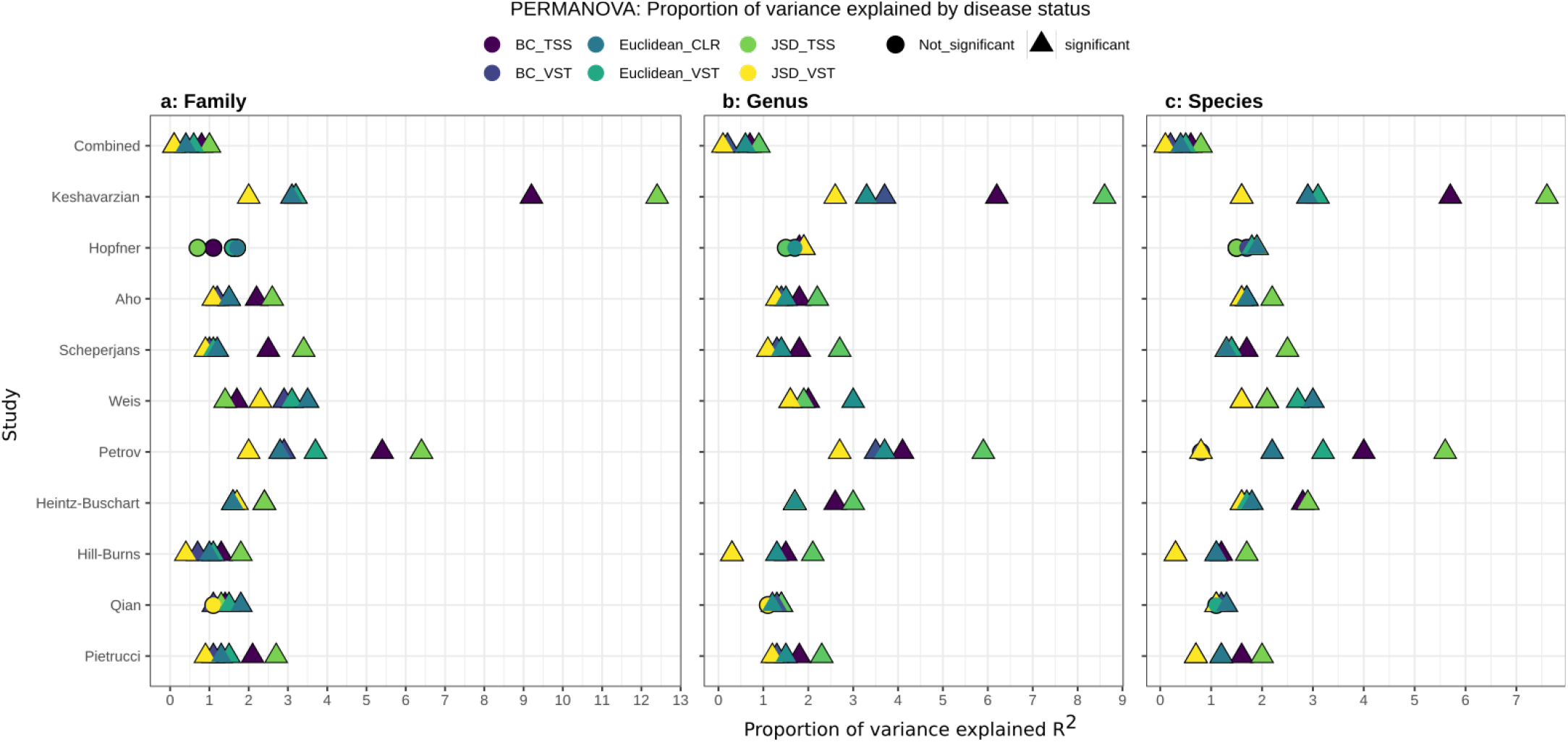
The gut microbiome structure differs significantly between PD patients and controls. Data were normalized using three independent approaches (Variance Stabilizing Transformation, VST; Total Sum Scaling, TSS; Centered Log-Ratio, CLR) and beta-diversity was estimated using three indices (Bray-Curtis, BC; Jensen-Shannon divergence, JSD; Euclidean). The effect of the disease status on the clustering of the data was assessed using a permutational analysis of variance (PERMANOVA). In the majority of the studies and approaches considered, and across all taxonomic ranks (a, b, c), the gut microbiota of PD patients resulted significantly different from the one of controls. The disease status explains only a small fraction of the data variability (< 13% R^2^), indicating that other environmental factors might have a stronger role in shaping the bacterial communities. The dataset obtained by pooling all ten studies is referred to as “Combined” in the figure.

We wanted to verify whether underlying differences, unrelated to the origin of the sampling cohorts, existed between the gut microbiome of PD patients and controls. Moreover, we aimed at identifying which study-specific factors most defined the differences across datasets. We used the normalization-distance pairs which best captured the variability of the data (Fig 3) to perform a distance-based Redundancy Analysis (dbRDA) on the pooled data (Supplementary Figures 4-6). First, we ordinated the combined data without constraints and without accounting for the variability introduced by the study. In accordance with the previous PERMANOVA analyses, the separation of the samples was driven by the study of origin (Supplementary Figures 4-6). We then inferred the degree of similarities between studies using the sample coordinates in the dbRDA (Supplementary Figures 7, see methods for details). The only four strongly divergent datasets were from: Weis et al.^24^, who used the sequencing platform IonTorrent; Hopfner et al.^14^, who maintained the samples at room temperature up to 48 h and analyzed them using the V1-V2 variable region of the 16S rRNA gene; Keshavarzian et al.^11^ who collected samples in anaerobic pouches; and from Heintz-Buschart et al.^13^ who immediately flash-froze the samples after collection and used a lab-specific DNA extraction protocol. We then verified which study-specific aspect (e.g. sequencing strategy, DNA extraction) most influenced the structure of the bacterial communities. We created additional dbRDA constraining the data by each potential confounding factor. Each factor significantly shaped the clustering of the data when considered individually, and this was observed for all taxonomic ranks and normalization approaches we used (Supplementary Table 2). In general DNA extraction protocols, country of origin, and 16S variable region were the factors that explained most of the variance within the data (Supplementary Table 2). Finally, we compared the dbRDA models constrained by all confounding factors and disease status with the one constrained only by the disease status and study and verified that both models explained the same proportion of data variability (Supplementary Table 2). Hence, removing the influence of the study will simultaneously eliminate the effects of other known study-specific confounding factors.

Accounting for the variability introduced by the study within the dbRDA drastically decreased the batch effect, irrespectively of the normalization-distance pair used (Supplementary Figures 4-6). However, samples did not cluster according to the disease status, suggesting that the environmental variability is higher than the variability explainable by the disease. Therefore, we constrained the dbRDA conditioned for study by disease status, to maximize the divergence between PD and control samples. We used this approach to determine the main taxa driving the separation between conditions. In accordance with the above results, the newly created constrained axis along which PD and controls diverged, significantly explained the clustering of the data (ANOVA-like permutation test; Fig 4, Supplementary Figure 5-6), but accounted for only <1% of the data variability. Since the constrained ordination obtained for the TSS-JSD pair explained a slightly higher proportion of variance, we selected this approach to identify taxa that strongly influenced the separation of the samples. The divergence between PD and controls was mainly driven by the family Bifidobacteriaceae and Akkermansiaceae, which were more enriched in PD, and the family Lachnospiraceae, which was more abundant in control samples (Supplementary Figure 8). Similarly relevant, but with a minor difference between conditions were the families Rikenellaeae, Porphyromonadaceae, Christensenellaceae, and the *Clostridium methylpentosum* group in the Oscillispirales order, all of which were more enriched in PD (Supplementary Figure 8). These results were mirrored in the dbRDA performed using genus and species abundances, which revealed that species in the *Akkermansia* and *Bifidobacterium* genera were strongly enriched in PD, whereas several species belonging to the Lachnospiraceae family caused the divergence of control samples (Supplementary Figure 7, Fig 4). When we omitted the dataset of Schepejans et al^15^, the overall results did not change, and only minor differences were observed (some genera and species in the Lachnospiraceae and Christensenellaceae family were not detected as main drivers of sample separation).

**Figure 4.**
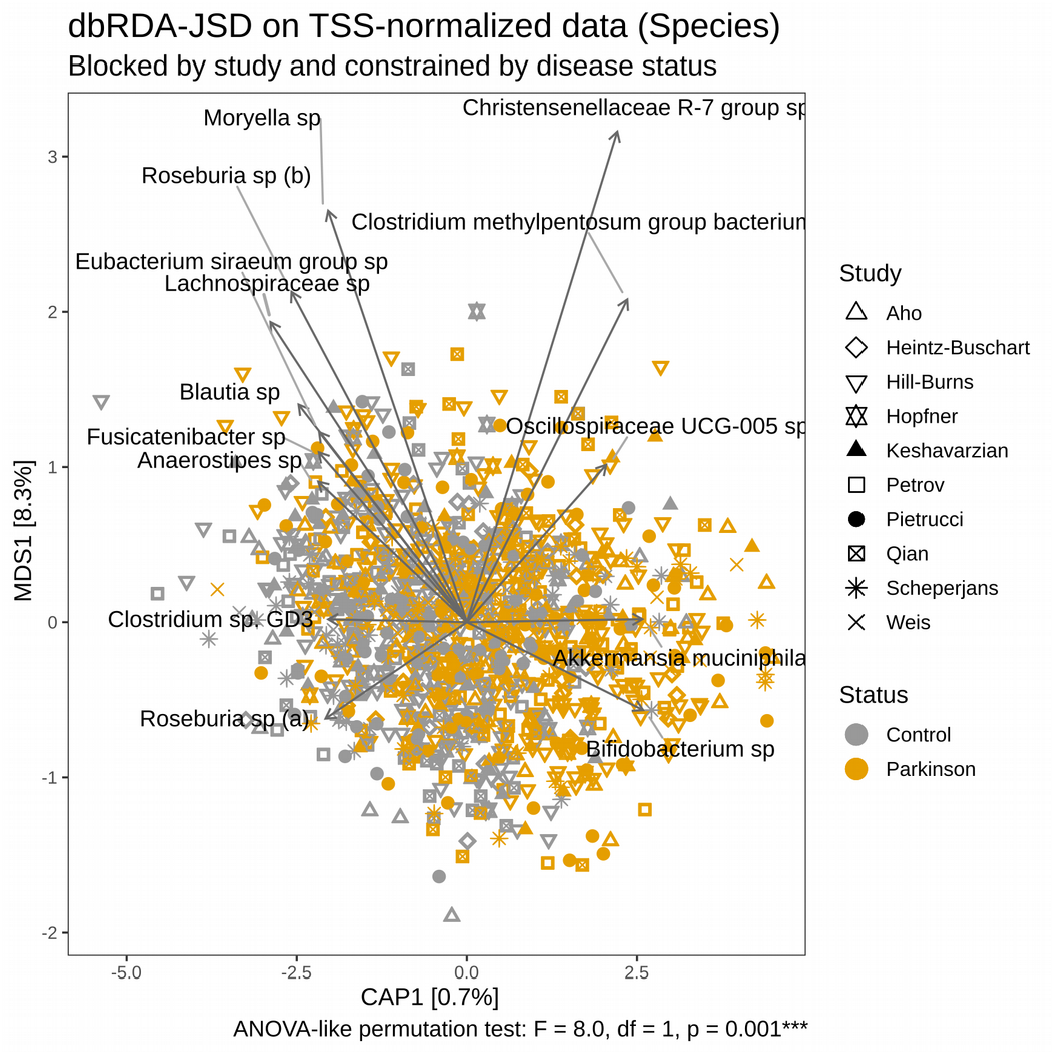
Most important species driving the divergence of the gut microbiota between PD patients and controls. Distance-based Redundancy analysis (dbRDA) was performed on Jensen-Shannon divergence (JSD) calculated on data normalized through total sum scaling (TSS). dbRDA was conditioned by study and constrained by disease status. Data refer to species abundances. The limited proportion of data variability explained by the axis constrained for disease status (CAP1) indicates that environmental factors have a major influence in shaping the bacterial communities. However, the influence of the disease status on the community structure is statistically significant (ANOVA-like permutation test). Only taxa showing a significant association with the clustering of the samples and the strongest abundance variation between the conditions are reported.

### The gut microbiome of PD patients and controls are enriched in different bacterial taxa

In the first instance, we analyzed all ten datasets individually using three separate approaches. The number of taxa that showed a statistically significant difference in abundance between PD and controls varied greatly across the studies and methods we used (Supplementary Figure 9, Supplementary Table 3). Amongst all studies, the highest number of significant taxa were detected in the datasets of Hill-Burns et al.^12^ and Petrov et al.^17^, whereas the lowest number was observed in Qian et al.^18^ and Hopfner et al.^14^ (Supplementary Figure 9, Supplementary Table 3). To obtain a generalizable overview of the PD-associated microbiome, we combined two independent approaches that we refer to as Pooled data and Pooled results approach. In the first, we pooled the count tables obtained for each study and used the same three methods we applied to the individual datasets and statistically accounted for the variability introduced by the study. Taxa were considered differentially abundant between PD and controls when detected as statistically significantly different by two out of three methods. This first list of taxa was then merged with the outcome of the Pooled results approach, in which we first estimated the differences in abundance for each taxon in the individual datasets and then used random-effect meta-analysis to pool the results (Fig 5, Supplementary Figures 10, 11, Supplementary Table 4).

**Figure 5.**
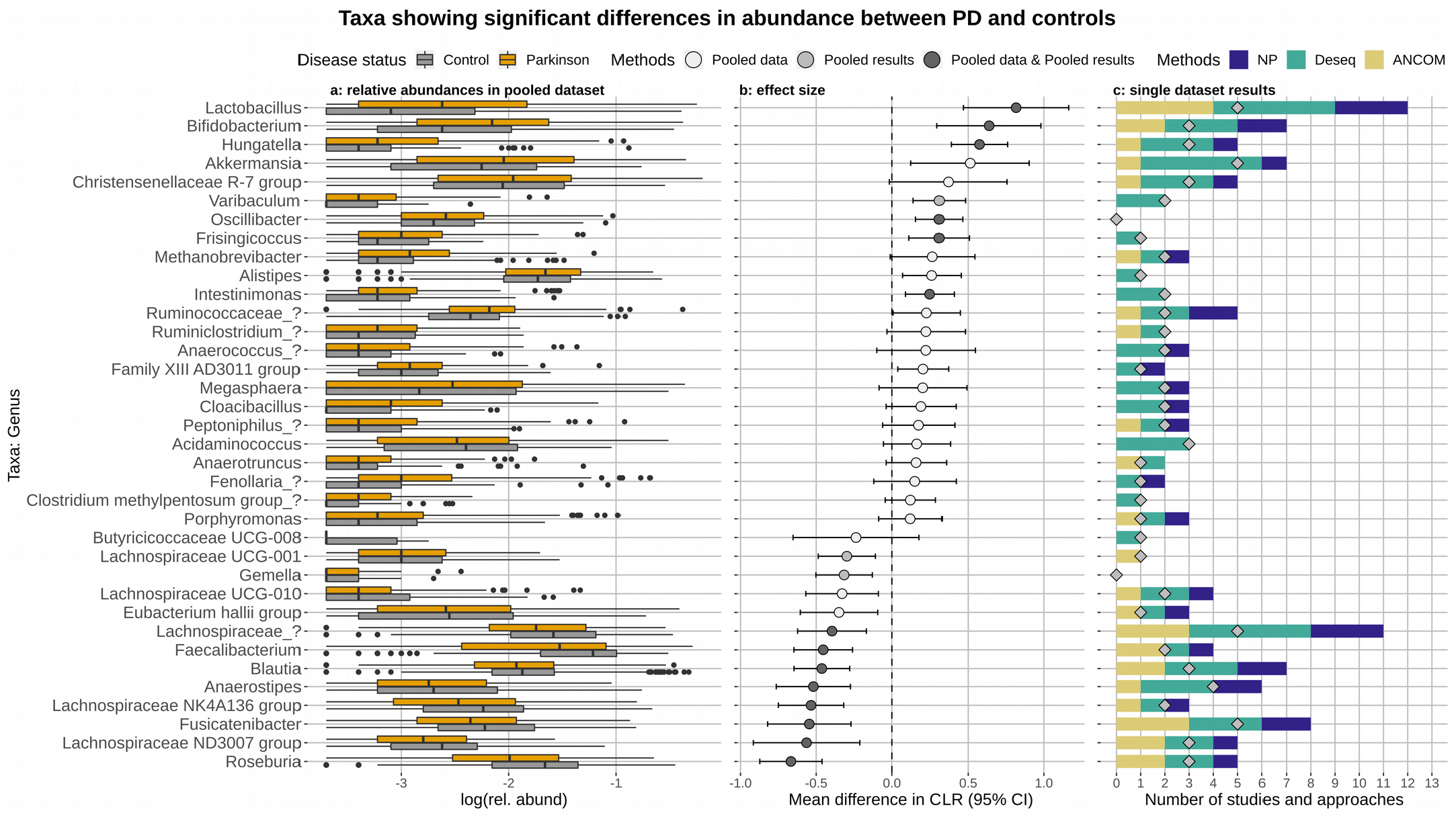
Genera showing a significant difference in abundance between PD patients and controls. The relative abundances of the genera retrieved from the pooled data are reported in panel a. Effect size was estimated via the mean difference in CLR (b) using a random-effect meta-analysis approach on all taxa resulting differentially abundant in the Pooled results or Pooled data approaches. The color of the dots indicate which of the above two approaches detected the taxa differentially abundant. Taxa more abundant in controls have an effect size shifted to the left, whereas taxa more abundant in PD have an effect size shifted to the right. C shows the number of times each genus was detected differentially abundant between PD patients and control samples across studies (diamonds) and approaches (bars). We used ten studies and three approaches, hence the maximum number of times a taxon can be detected differentially abundant is 30.

After obtaining a first list of differentially abundant taxa, we used the metadata made available by five of the ten studies we re-analyzed to verify whether age and/or gender influenced the abundances of the taxa we detected to be enriched or depleted in PD. By comparing generalized linear mixed models (GLMMs) with and without accounting for the disease status, we identified taxa that showed a significant alteration in abundance in PD irrespective of age and gender (Supplementary Table 5). For most of the taxa, the disease status was required to best explain taxa abundances, underlining the robustness of our approaches. We did not detect any taxa for which gender and/or age alone could best describe the data. However, for 11 Species, 13 Genera, and 7Families the available data did not allow to clearly establish whether the disease status was an essential factor to explain taxa abundances (Supplementary Table 5). Hence, we did not consider these taxa further. For example, the Prevotellaceae family and a species within this family (Supplementary Table 5) were both detected differentially abundant between PD and controls. However, the abundance of both taxa was also influenced by gender and age (Supplementary Table 5, Supplementary Figure 12).

Among the remaining genera, after controlling for available metadata, *Roseburia, Blautia, Fusicatenibacter, Faecalibacterium, Anaeorostipes* and other two unknown genera belonging to the family Lachnospiraceae were strongly enriched in the control samples (Fig 5B) in several datasets (Fig 5C). Consistently, the Lachnospiraceae family and species within this family and affiliated to the *Fusicatenibacter, Blautia*, and *Roseburia* genera were strongly enriched in control samples (Supplementary Figures 10, 11). The family Butyricicoccaceae was also more abundant in controls, even though it was detected differentially abundant in fewer studies (Supplementary Figure 10). PD samples were instead most often enriched in the genera *Lactobacillus, Bifidobacterium, Hungatella*, and *Akkermansia* (Fig 5). Additionally, the R-7 group of the Christensenellaceae family, the genera *Methanobrevibacter, Oscillobacter, Frisingicoccus and Varibaculum* were also detected more abundant in PD, but with a smaller effect size (Fig 5). PD samples were enriched in species belonging to different taxonomic groups e.g. Ruminococcaceae, Christensenellaceae, *Bifidobacterium, Lactobacillus, Hungatella*, and *Alistipes* (Supplementary Figure 11). Other species, such as *Intestinimonas* sp, *Oscillibacter* sp also resulted more abundant in PD, but in fewer studies (Supplementary Figure 12). In contrast, the majority of species enriched in controls belonged to the families Lachnospiraceae and Ruminococcaceae (Supplementary Figure 12). The shifts in taxa abundances outlined above were robust to the sensitivity analysis we performed omitting the baseline data of the longitudinal Finnish cohort, which resulted in minor differences affecting only taxa having a small effect size (Supplementary Figure 13).

Finally, for each dataset, we obtained hypothetical functional prediction based on the 16S profiles. Differential abundance testing of predicted-pathways between PD and controls was performed as for the taxonomic data. The majority of predicted-pathways enriched in PD were related to ubiquinone (Coenzyme Q; CoQ) and menaquinone (vitamin K2) biosynthesis. 4-aminobutanoate (GABA) degradation and glutamine-glutamate metabolism, methanogenesis, and lactic-type fermentation (Supplementary Table 3, 4; Fig 6, Supplementary figure 14). Instead, the control samples were enriched in pathways involved in the biosynthesis of cobalamin (vitamin B12), degradation of glucuronate and galactoglucuronate, and methane production from acetate degradation (Supplementary Table 3, 4; Fig 6, Supplementary Figure 14).

**Figure 6.**
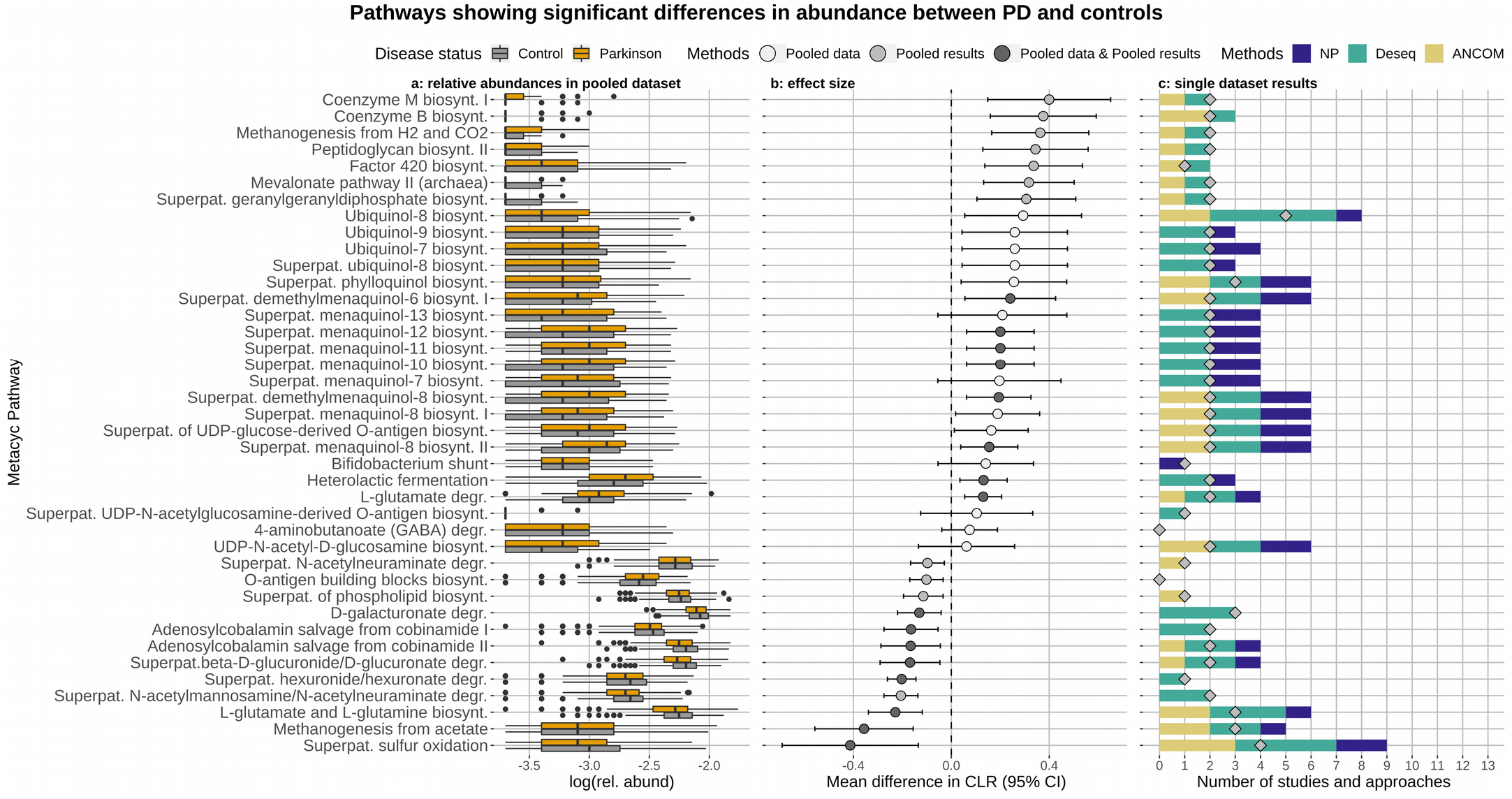
Pathways showing a significant difference in abundance between PD patients and controls. Only selected relevant pathways are shown (a full overview is reported in Supplementary Figure 13). The relative abundances of the pathways retrieved from the pooled data are reported in panel a. Effect size was estimated via the mean difference in CLR (b) using a random-effect meta-analysis approach on all taxa resulting differentially abundant in the Pooled results or Pooled data approaches. The color of the dots indicate which of the above two approaches detected the pathway differentially abundant. Pathways more abundant in controls have an effect size shifted to the left, whereas pathways more abundant in PD have an effect size shifted to the right. C shows the number of times each genus was detected differentially abundant between PD patients and control across studies (diamonds) and approaches (bars). We used ten studies and three approaches, hence the maximum number of times a taxon can be detected differentially abundant is 30.

## Discussion

In recent years, several studies have analyzed the gut microbiome of PD patients and reported various degrees of alteration compared to healthy controls. We wanted to verify whether consistent changes in the gut microbiome of PD patients can be identified across studies, as often contrasting results have been reported. Hence, we performed a meta-analysis using all publicly available 16S rRNA gene amplicon datasets comparing PD patients with healthy controls, to address reproducibility and identify targets to foster further experimental work. By integrating different statistical methods we re-analyze all data and offer a comprehensive and robust consensus of the most consistent features of the gut microbiome associated with PD. Studies were heterogeneous in sampling populations and methodological approaches (Table 1, Supplementary Table 2; Supplementary Figure 7), and the inter-study variability was the main factor driving bacterial community structures (Fig 3, Supplementary Figures 4-7; Table 1, Supplementary Table 2), as observed in previous microbiome-based meta-analyses performed in the context of diet and colorectal cancer^27,28^. Our analyses suggest that particular methodological approaches, such as sample collection and transport, sequencing platform, and the chosen 16S variable region might be the main reasons for the heterogeneity across the datasets we considered (Supplementary Figure 7, Table 1, Supplementary Table 2). Variability across the studies was also a reflection of the different sampling populations used in the different studies (Table 1). For example, the proportion of PD patients using medication as well as the duration of the disease varied across datasets. Also, controls were sometimes closely matched but sometimes differed considerably from cases in terms of age, sex and ethnicity. Hence it is important to stress that using the available data it is impossible to determine whether the associations reported are causally linked to PD.

By stratifying the analysis by study, we could simultaneously exclude the effect of all other known study-specific confounding aspects, such as country of origin, DNA extraction, and sequencing (Supplementary Table 2, Supplementary Figures 4-6). In agreement with previous studies^11-13,15,18,19^, we show that the gut microbiome of PD patients significantly differs from the one of controls (Fig 3, Supplementary Figure 8). Although PD can explain only a limited portion of data variability (Fig 3, Supplementary Figures 4-6; Supplementary Table 2) the observed differences are robust to technical confounders and across sampling cohorts, indicating that the alteration in the gut microbiome of PD patients is a general phenomenon. The analysis of the bacterial alpha diversity suggests that such alterations might be explained by a decrease in the abundances of the most abundant species and an increase in the rare ones (Fig 1, Supplementary Figure 1), which is a typical alteration observed in dysbiosis associated with inflammatory bowel disease and irritable bowel syndromes (IBD, IBS)^29^. An increase in bacterial diversity in the gut microbiome of PD patients has been previously reported both in studies we re-analyzed and studies for which data were not available^11,18,21,30^. Similarly, a recent work reported no differences in OTU-based alpha-diversity but found that in controls 98% of OTUs could be assigned to the four dominant Phyla, whereas only 88% of OTUs belonged to these Phyla in PD^31^. This suggests a decrease in dominant taxa and an increase in less abundant ones as underlined by our results.

PD samples had a lower abundance of the genera *Roseburia*, *Fusicatenibacter*, *Blautia, Anaerostipes* (Lachnospiraceae family), and *Faecalibacterium* (Ruminococcaceae family) (Fig 5, Supplementary Figures 10-12; Supplementary Tables 3, 4), a finding seen in other neuro-inflammatory and rheumatologic disorders^32-34^. Most of these taxa are abundant and widespread bacteria in the gut microbiota of healthy individuals, they are major butyrate-producers and have often been found depleted in IBD^29^. Similarly depleted in IBD are bacteria of the Butyricicoccaceae family^35^, which are important butyrate producers and were highly depleted in PD in our analyses (Supplementary Figure 10). The depletion of these taxa suggests a low level of butyrate in the gut of PD patients. Butyrate is a fundamental energy source for intestinal epithelial cells and has been reported to reinforce the intestinal epithelium as well as preventing inflammation and carcinogenesis^36^. Our findings are consistent with previous studies showing low levels of butyrate and increased gut permeability and inflammation in PD patients^7,9^. SCFA are not only relevant for gut health, but they can also influence the ENS, have systemic anti-inflammatory properties, promote normal microglia development, and potentially affect epigenesis in the CNS^3^. Importantly, PD patients have been shown to have increased levels of various cytokines in both the colon and serum, suggesting that they suffer from systemic inflammation which could result in microglial activation driving disease progression^37^. Altogether these data suggest that the alteration in SCFA production in the gut combined with an increase gut permeability and inflammation might have systemic implications in PD. To the best of our knowledge, only Aho et al^16^ identified bacteria of the Butyriciccocaceae enriched in controls. Similarly, only Aho et al^16^ and Weis et al^24^ detected *Fusicatenibacter* to be significantly depleted in PD patients. Specifically, Weis et al. report that the decrease of this genus together with *Faecalibacterium* was correlated to the degree of gut inflammation^24^. Interestingly, both these genera were low in abundance in IBS and ulcerative colitis^38,39^, and *Faecalibacterium* showed strong anti-inflammatory and protective effects in an acute colitis mouse models^40^. Our analysis suggests that the depletion of taxa playing a key role in maintaining gut health is widespread in PD across populations. Such depletion resembles dysbiosis observed in other gastrointestinal dysfunctions (e.g. IBD) and supports the link between PD and gut health as underlined by retrospective studies indicating that the overall risk of developing PD in IBD was significantly higher, reaching 28% and 30% increase in patients with Crohn’s and ulcerative colitis, respectively^37^.

Our results indicate a higher abundance of the genera *Lactobacillus*, *Akkermansia*, *Hungatella*, and *Bifidobacterium* in PD gut microbiome (Fig 5, Supplementary Figures 10, 11; Supplementary Table 3, 4). The fact that the more abundant genera in PD belong to different families and even orders, support the idea of increased diversity in the gut-microbiota of PD patients (Fig 1, Supplementary Figure 1). The genus *Lactobacillus*, and the Lactobacillaceae family, were the most strongly enriched taxa in PD across the studies we re-analyzed, in line with previous findings^11,12,17,19,41^. *Lactobacillus* strains are low abundant members of the gut microbiota and their abundance varies greatly across human disease and chronic conditions^42^. Some strains of *Lactobacillus* and *Enterococcus*, also enriched in PD (Supplementary Figure 11), are able to produce enzymes that can degrade levodopa into dopamine, suggesting that their abundances might be a consequence of the use of this medication in PD^43,44^. Levodopa is absorbed in the small intestine, but it has been reported that 10-20% can reach the large intestine^45^, indicating that a substantial amount of this molecule can be readily available for gut bacteria and could help these bacteria to proliferate. Interestingly, none of the Chinese studies conducted so far detected *Lactobacillus* or the family Lactobacillaceae enriched in PD^18,20,21,23,46^. Consistently, these taxa were not enriched in PD samples in the only Chinese study included in our meta-analysis. The majority of *Lactobacillus* species have been found in food (e.g. dairy, milk, fermented food, probiotics)^42^. Hence, it is possible that differences in diet between regions could explain the difference between the Chinese cohorts and the others.

*Akkermansia* has been repeatedly shown to be more abundant in PD compared to controls^11-13,20,22^. In general *Akkermansia* spp. are considered beneficial for human health, as they fortify the integrity of the epithelial cell layer and can modulate the immune system^47,48^. For example, a recent study reported that these bacteria ameliorate age-related decline in colonic mucus thickness and attenuate immune activation in accelerated aging mice^49^. However, contrasting results regarding the influence of *Akkermansia* spp. on gut health exist^50^. Intriguingly, constipated individuals have been shown to have a gut microbiome enriched in *Akkermansia*^51-53^, and constipation is one of the major non-motor symptoms in PD, often starting decades before motor symptoms arise. The increase in *Akkermansia* could be a consequence of constipation, even though animal studies suggest that this genus might contribute to an increased transit time. *Akkermansia* spp. are mucin-degrading bacteria and they can lead to a depletion of the intestinal mucus-layer when the gut microbiota is unbalanced^54,55^. Mice receiving stool from chronically constipated patients showed drier stools, decreased number of goblet cells, and impaired intestinal barrier function in association with an increase in *Akkermansia* spp^54^. The unbalanced microbiota observed in PD patients, might lead to a proliferation of *Akkermansia* spp, which in turn might lead to decreased mucus thicknesses, drier stools, and constipation. It is important to point out that multiple strains belonging to the same *Akkermansia* species can co-exist in the gut and the modulation of host-response can be strain-specific^56,57^. For example, different *Akkermansia munichipalia* strains have different effects on the differentiation of Regulatory T cells (Tregs) and SCFA production^57^, both factors altered in blood and gut, respectively, of PD patients^6,9,58^. Altogether these data indicate that the increased abundance of *Akkermansia* spp in PD might be linked to alterations in the immune response and constipation. These effects might be strain-specific and more in-depth strain-resolved metagenomics are needed to elucidate these aspects in PD.

Among the most abundant taxa in PD there were bacteria belonging to the Christensenellaceae family (Fig 5, Supplementary Figure 10, 11), in line with previous reports^11,12,21,41^. This family is widespread in the gut of the human population and it is generally associated with healthy phenotypes, even though their abundances positively correlate to the intestinal transit time^59^. *Christensenella* spp. can efficiently support the proliferation of *Methanobrevibacter smithii* via H_2_ production, explaining the recurrent co-occurrence between these two taxa^60^. *Methanobrevibacter* belongs to the Archaea domain and it is the major hydrogenotrophic methane producer in the human gut, and was also more abundant in the PD samples we re-analyzed (Fig 5). In both controls and PD patients the abundances of these taxa were positively correlated (Spearman rank test: PD, Z = 10.3, p-value 0.0005; controls, Z = 8.8, p-value 0.0005; Supplementary Figure 15). Moreover, the 16S-based functional prediction we performed showed that the pathways for the formation of methane from H_2_ and CO_2_ and for the synthesis of key co-factors involved in methanogenesis (Coenzyme B and M, Factor 420) are strongly enriched in PD (Fig 6). The increased abundance of these two co-occurring genera in PD patients might contribute to the production of methane, which in turn could influence the intestinal transit. In fact, *Methanobrevibacter* is enriched in constipated patients^52 53^, just as *Akkermansia*, and growing evidence indicates that methane decreases peristaltic movements slowing down intestinal transit time^61^. Surprisingly, to the best of our knowledge, only one other study reported an enrichment in *Methanobrevibacter* in PD^20^. It is worth noting that in the 16S-based functional prediction another pathway that produces methane through the degradation of acetate was enriched in controls (Fig 6). This pathway is mainly found in Archaea of the genus *Methanosarcina*. However, we did not detect these taxa enriched in controls. They were identified as significantly more abundant in controls only in the dataset of Qian et al^18^ and only by a single method (Supplementary Table 3). Hence, these data need to be interpreted with caution as they might be an artifact of the 16S-based predictions. It is important to specify that the abundance of Archaea in the human gut microbiota is considerably lower than that of Bacteria, and current methodologies (DNA extraction, primers used for 16S amplification) strongly discriminate against Archaea^62^. Hence, it is possible that the abundances and the diversity of these microorganisms is currently poorly represented in the available datasets.

In agreement with the only shot-gun metagenomic study so far available^10^, we identified several predicted-pathways involved in galacturonate and glucuronate degradation depleted in the PD-microbiome (Fig 6, Supplementary Figure 14). The enrichment of GABA degradation pathways and glutamate/glutamine biosynthesis pathways in PD suggests an alteration in the enteric production of these neurotransmitters. The gut microbiota has been previously suggested to alter the glutamate-glutamine-GABA cycles in schizophrenia and autism^63,64^, and alterations in the level of this transmitter have been found in brains of PD patients^65,66^. Hence, it is intriguing to speculate that the gut microbiota might play a role in modulating these chemicals in PD patients. Further experimental work will be required to verify whether these metabolic changes in the PD microbiota can induce alterations in the CNS. Surprisingly, the majority of the predicted-pathways enriched in the PD microbiota were related to ubiquinone (CoQ) and menaquinone (vitamin K2) biosynthesis. Data from animal and pre-clinical studies showed that both CoQ and vitamin K have a crucial role in avoiding the mitochondrial dysfunctions observed in PD^67,68^. Hence, the increased biosynthetic capacity we observed in the PD-associated microbiota is surprising. Although these findings would need to be confirmed via e.g. shot-gun metagenomics/metabolomics, it is tempting to speculate that the potential increase of vitamin K2 production in the gut might increase systemic concentrations of these chemicals in PD. Interestingly, vitamin K plays an important role in the biosynthesis of sphingolipids^69^ which are emerging as an important determinant in PD development^70^. These data suggest novel mechanisms through which the gut microbiota might potentially influence PD development.

In summary, our analyses reveal underlying consistent differences in the average gut microbiota composition between PD patients and controls. Variations among studies are the strongest factor in shaping the data structure, but by accounting for the variability derived by the sampling cohorts we were able to show that the alteration of the gut microbiome in PD is consistent across studies and countries. The differences in taxa abundances between PD and controls indicate that the gut microbiota of PD patients shares similarities with those of other neurological and inflammatory gastrointestinal diseases. Taxa important in maintaining gut integrity and health via the production of SCFA are depleted in PD and this together with the growing evidence of gut and systemic inflammation in PD, point towards an important role of the gut microbiota in modulating the immune function in this disease. Moreover, we were able to identify previously overlooked taxa enriched in PD such as *Methanobrevibacter*, Butyriciccocaceae, and identified some potentially new metabolic routes through which the microbiota might influence PD. Our findings align with the accumulating evidence indicating gut and systemic inflammation in PD, and suggest that the dysbiotic gut microbiota could influence host immune function and be linked to the gastrointestinal symptoms often recurring in PD patients.

## Materials and Methods

### Study selection

#### Search strategy

On the 29^th^ of March 2020 Google Scholar was searched for publications that contained all the words “16S”, “gut”, “Parkinson”, “metagenomic”, the exact phrase “Parkinson’s disease”, at least one of the words “microbiota” [OR] “microbiome” [OR] “gut” [OR] “intestinal” anywhere in the article. This resulted in 1,010 entries. Titles were then manually screened and if they contained the words “microbiome” or “microbiota” and “Parkinson’s disease” the abstracts were further consulted. Moreover, the Sequence Read Archive (SRA) in NBCI was queried with the following term “Parkinson” [AND] “microbiome”, resulting in two additional studies (Bioprojects): PRJNA530401 and PRJEB14928. We managed to match only the latter Bioproject ID to a published study^14^, hence we considered only this dataset in our analyses.

#### Inclusion criteria

We included all studies comparing the composition of the gut microbiota between patients with confirmed PD to a control population without PD, and that made the raw reads of the 16S rRNA gene amplicon sequencing available. Studies with any design (e.g. cohort studies, case-control studies, or cross-sectional studies), and from any geographical area were included. Studies could use any method for the acquisition and analysis of samples. We identified a total of 23 studies that cataloged the gut microbiome of PD patients using metagenomics (Supplementary Table 1). 9 of these studies did not make the raw data publicly available. We were unsuccessful in obtaining the raw reads from the authors, as data were either protected by ethical restriction or the authors did not answer our requests. In other cases the raw reads were available, but it was impossible to associate the data with the disease status as this information was not reported in the metadata. The samples from Scheparjans et al^15^, originally sequenced using a 454 technology, were recently re-sequenced in a follow-up study by the same group using Illumina MiSeq^16^. Hence, we only included in our analysis the most recent datasets. Finally, one study used shot-gun metagenomics^10^ and three studies were available only as pre-prints at the time of writing and the raw reads were not made public yet.

### Data retrieval and zOTU picking

Raw reads were downloaded from SRA or the European Nucleotide Archive (ENA). Adapters were removed using the bbtools suit^71^. Data were analyzed using Lotus^72^ and the UNOISE3^73^ algorithm for zOTUs calculation, bundled in a new Lotus version (Lotus2), currently under development. Due to the technical variability among datasets (e.g. 16S region, sequencing technology) the filtering parameters used by the *sdm* program called by Lotus, were adjusted for each dataset independently and are reported in the supplementary materials (Supplementary Table 6). For the datasets of Petrov et al^17^ and Weis et al^24^ we had to decrease the accepted minimum error due to the low quality of the sequencing data (Supplementary Table 6). 16S-based functional predictions were obtained using the default settings in *picrust2* v2.3.0-b^74^ and the Metacyc database. In this analysis, the dataset of Qian et al.^18^ was not included, as with the default cutoffs the sequences aligned poorly with the reference database used. Count tables for species, genera, families, and functional predictions were then analyzed using R v3.6.2^75^. Datasets were processed using the *phyloseq* R-package^76^, samples with < 4,500 reads were removed, and taxa with < 5 counts and predicted-functionalities with < 20 counts in < 2.5% of samples were removed. This filtration steps left a total of 1,211 (530 control, and 681 PD samples) and 1,121 samples (485 control and 636 PD samples) for the taxonomic and predicted-function data, respectively. Finally, enterotypes were predicted using rarefied relative abundances of genera via the https://enterotypes.org/web-platform.

### Statistical analyses of single studies

#### Analysis of alpha-diversity

Alpha-diversity indices at the species level were calculated using the *microbiome* R-package^77^, after rarefying without re-sampling at the even depth of 5,000. Due to rarefaction 8 samples were further removed, leaving a total of 1,203 samples (523 control and 680 PD samples). We measured richness using the number of Observed species, the Chao1, Fisher’s alpha, and ACE indices; evenness using the Bulla and Simpson indices, dominance using the core abundance, which measures the relative proportion of core species that exceed relative abundance of 0.2% in over 50%, and the Simpson’s index of dominance. Finally, we estimated rarity using the low abundance index, which considers the relative proportion of the least abundant species below a detection level of 0.2%, and the rare abundance index, which estimates the relative proportion of the non-core species exceeding the detection level of 0.2% at 50% prevalence. Additionally, we calculated the ratios of Firmicutes to Bacteroidetes phyla and *Prevotella* to *Bacteroides* genera, as log_2_ ratios of their relative abundances. In each dataset, the differences in alpha-diversity between control and PD samples were assessed using Agresti’s generalized odd ratios using the *genodds* function in the *genodds* R package^78^. This statistic, based on ranks and analogous to the U statistic underlying the Mann-Whitney test, does not make strong assumptions about the distributions of measures and is comparable between measures of diversity with different scales.

#### Beta-diversity and differential abundance analyses

For each dataset, beta-diversity and differential abundance analyses were performed using three independent approaches (described in the sections below): i) normalization via total sum scaling (TSS; i.e. relative abundances) and differential abundance (DA) inference through Wilcoxon-Mann-Whitney (WMW) tests; ii) variance stabilizing transformation (VST) and DA inference using DESeq2^79^; iii) compositional approach based on centered log-ratios (CLR) and DA inference using analysis of composition of microbiomes (ANCOM)^80^. We then reported the number of times each taxon showed a significant difference in abundance between PD and controls across studies and statistical approaches. For example, a taxon detected differentially abundant across all ten datasets and all three approaches would have a final score of 30 (panel C in Fig 5, Supplementary Figures 10, 11, 14). Differential abundances of *picrust2* predicted functionalities between PD and controls were inferred using the same approach outlined above. The rarefaction used in the TSS approach did not result in a loss of samples for the 16S-based predicted functionalities.

#### Total sum scaling (TSS) and non-parametric tests

After rarefying without re-sampling at the even depth of 5,000, data were normalized by dividing the counts of each taxon for the total counts of all taxa (total sum) in the sample. Beta-diversity matrices were calculated using the Bray-Curtis (BC) dissimilarity index and the Jensen-Shannon distances (JSD). Statistical differences between control and PD groups were then tested using the permutational multivariate analysis of variance (PERMANOVA) as implemented in the *adonis2* (analysis of variance using distance matrices, ADONIS) function in the *vegan* R-package^81^. DA analysis was performed using a two-sided WMW test, using Benjamini Hochberg (BH) p-value correction.

#### Variance stabilizing transformation (VST) and DESeq2 analyses

Since the DESeq2 approach does not account for zero-inflated data, the correction factors were calculated using the GMPR method that is based on geometric means of pairwise ratios^82^. Euclidean, BC, and JSD distances were used as beta-diversity estimators after normalizing the data via VST. Statistical differences between control and PD groups were tested using the *adonis2* function as specified above. DAs were calculated using default DESeq2 parameters that include a negative binomial GLM fitting and a Wald test^79^. Multiple testings were accounted for using BH p-value correction.

#### Compositional analysis: Centered log-ratios (CLR) and ANCOM

Data were transformed using CLR, after imputing zeros through Bayesian-multiplicative replacements via the count zero multiplicative approach (“CZM”) in the *cmultRepl* function of the *zCompostions* R package^83^. Euclidean distances, that for such data correspond to Aitchison distances, were then calculated^80^. Statistical differences between control and PD groups were tested using the *adonis2* function as specified above. DA analysis was performed via the ANCOM approach as implemented in the R script *ancom_v2.0^Si^* using a 0.95 zero-cutoff and significance at the 0.6 percentile.

### Statistical analyses of the combined studies

#### Analysis of alpha-diversity

The Agresti’s generalized odd ratios estimated for each alpha-diversity index and each individual study were pooled using a random effect meta-analysis using the function *metagen* in the R package meta^85^.

#### Analysis of beta-diversity

Count tables obtained for each dataset were pooled and beta-diversity analyses were performed using the three approaches described above (TSS-, VST-, CLR-based analysis). For each normalization approach, statistical differences between control and PD groups and the marginal effects of study and disease status were tested using the *adonis2* function. We then used the distance measure that captured a highest fraction of the variability in the pooled dataset to compute distance-based Redundancy Analyses (dbRDA). dbRDAs were performed using the *“CAP”* option in *phyloseq*, which calls the *capscale* function in the *vegan* package. Data were clustered without conditioning (blocking) for studies and without constraining, by conditioning for study, and by conditioning for study and constraining for disease status (PD vs Control):

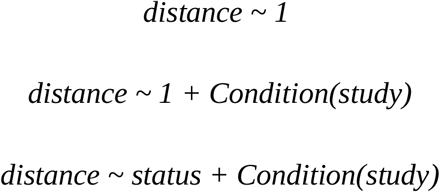

The significance of the constrain was tested using an ANOVA-like permutation test *(anova.cca* function in the *vegan* R package). For each normalization method, we investigated the effect of study-dependent confounding factors such as country, sequencing platform (e.g. MiSeq vs IonTorrent), sequencing approach (single-end vs paired-end), amplified region (e.g. V4 vs V1-V2), and extraction method and type by creating additional dbRDAs and constraining the data for each individual factors. The effect of each constraining variable was tested using an ANOVA-like permutation test. We then verified whether accounting for the variability introduced by the study alone will allow us to simultaneously account for the variation derived by the other technical confounding factors. We compared the adjusted R^2^ (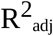) of a dbRDA obtained using the full model *distance ~ country + 16Sregion + ends + seq + extraction + extraction type + status* with the one of a reduced model including only disease status and study *(distance ~ study + status)*. Similar R^2^ _adj_, differences ≤ 0.1%, indicates that the two models are equivalent. The influence of confounding factors on microbial community structure was assessed at the Species, Genus, and Family level. Finally, we used the TSS normalized data to correlate the relative abundance of the taxa to the constrained and conditioned dbRDA via the *envfit* function in the *vegan* package. We selected only taxa significantly correlated with the clustering *(p-value* < 0.01), and showing the highest degree of variation (> |0.1| for genus and species, and > |0.075| for family) along the constrained axis (CAP1).

#### Similarity amongst studies

We used the unconstrained and unconditioned dbRDA performed on the TSS normalized species data to estimate dissimilarity amongst studies. We selected the coordinates of each sample across all axis that explained 90% of the data variance. These scores were then used to calculated Euclidean distances amongst samples. We then calculated distances between study centroids using the R package *usedist*^86^. Similarity among studies was then visualized using non-metric multidimensional scaling (NMDS) via the *metaMDS* function the in the *vegan* R package.

#### Differential abundance analysis

We combined two independent approaches to gather a consensus view on the taxa differentially abundant between PD patients and controls. We refer to these two approaches as Pooled data and Pooled results. In the Pooled data approach, the count tables obtained for each dataset were pooled and processed with the same methods used for the single datasets: i) TSS normalization on rarefied data and *independence_test* in the coin R package^87^ blocking data for the study; ii) DESeq2 approach adding the *“study”* variable as a covariate in the model; iii) ANCOM performed using a mixed-effect model with the effect of PD allowed to vary with study (via the formula *“random.formula = “~1+ status|study”)*, using a zero-cutoff 0.975 and significance at the 0.6 percentile. For all three methods BH p-value correction was used and the threshold for significance was set at ≤ 0.05. If a taxon or pathway had a significant difference in abundance in 2 out of three approaches, it was then retained (Consensus).

To this first list of differentially abundant taxa/pathways we added the data obtained from the Pooled results approach. In this approach, we normalized the count table of each individual dataset using CLR after adding a pseudo-count of 1 to 0 values. We then selected all taxa and pathways detected in at least 3 studies and estimated their shift in abundance between PD and controls using linear models for family, genera, and 16S-based predicted functionalities and Agresti’ generalized odd ratios for species. We then pooled these results using a random-effect meta-analysis via the *metagen* R function. The resulting p-values were corrected using BH. All taxa/pathways showing an adjusted p-value ≤ 0.05 and a 95% confidence interval (CI) not crossing 0 were retained.

Taxa and pathways showing significant differences in abundance between PD and controls in the Pooled data (2 out of 3 methods referred to as Consensus) or Pooled results approach were further considered. All taxa having abundances potentially influenced by age and/or gender were then removed (see below). For each taxa/pathway the effect size and the respective 95% CI were estimated using the Pooled results approach. Finally, correlation between the genera Christensenellaceae R-7 group and *Methanobrevibacter* was calculated on the relative abundances of non-rarefied data using a Spearman correlation test by blocking the data by study *(spearman_test* in the *coin* R package).

#### Influence of confounding factors on differential abundances

Of the ten studies we used only five had metadata available, and among the latter, only age and gender were always reported. We assessed the influence of age and gender on the abundance of the taxa we previously identified as significantly enriched or depleted in PD using generalized linear mixed models (GLMMs) controlling for zero-inflation as implemented in the R package glmmTMB^88^:

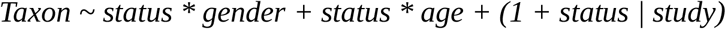

We created random slope and random intercept GLMMs for all taxonomic ranks we analyzed (species, genus, family). Models were fitted using either a negative binomial or a generalized Poisson distribution. First, we constructed zero-inflated and non zero-inflated models, and choose the best model using the Akaike information criterion (AIC; ΔAIC ≥ 2). We then created reduced models omitting each of the predictors (status, age, gender), their interactions (status:gender, status:age), and considering a constant effect of the disease status across studies (i.e. random effect = (1 *| study))*. We then compared all models using the *model.sel* function and the ΔAIC in the R package MuMIn^89^. If one of the best models (within a ΔAIC of 2) did not contain the variable disease status we concluded that the disease status might be not an essential factor needed to explain the taxon abundance. Hence, we removed these taxa from further analyses. If all best models contained the variable disease status, we consider PD as an essential factor shaping taxa abundances, thus we retained the taxa. For building the GLMMs, raw counts were used and data were rarefied to a fixed depth of 10,000 to avoid overparameterization.

## Data Availability

All data are attached to the manuscript

## Code availability

The R code used in this study will be made publicly available on GitHub after the peer-review process is completed.

## Acknowledgments

We are grateful for the technical support provided by A. Telatin and the assistance from S.J. Green and A. Keshavarzian in obtaining the raw reads of their study from SRA. This research was supported in part by the NBI Computing infrastructure for Science (CiS) group through access to High-Performance Computing infrastructures. The authors gratefully acknowledge the support of the Biotechnology and Biological Sciences Research Council (BBSRC); this research was funded by the BBSRC Institute Strategic Programme Gut Microbes and Health BB/R012490/1 and its constituent project BBS/E/F/000PR10356. GS was supported by the BBSRC Core Capability Grant BB/CCG1860/1. We acknowledge M. Mayer and R. Ansorge for constructive comments on the manuscript.

## Contributions

SR designed the study, performed bioinformatic and statistical analyses, interpreted the results and wrote the paper. GMS assisted in statistical analyses and data interpretation. JRB helped in data interpretation. FH provided support in bioinformatic and statistical analyses, and data interpretation. AN and IGC helped in data interpretation.

